# Validity of Wrist and Forehead Temperature in Temperature Screening in the General Population During the Outbreak of 2019 Novel Coronavirus: a prospective real-world study

**DOI:** 10.1101/2020.03.02.20030148

**Authors:** Ge Chen, Jiarong Xie, Guangli Dai, Peijun Zheng, Xiaqing Hu, Hongpeng Lu, Lei Xu, Xueqin Chen, Xiaomin Chen

## Abstract

**Aims:** Temperature screening is important in the population during the outbreak of 2019 Novel Coronavirus (COVID-19). This study aimed to compare the accuracy and precision of wrist and forehead temperature with tympanic temperature under different circumstances.

**Methods:** We performed a prospective observational study in a real-life population. We consecutively collected wrist and forehead temperatures in Celsius (°C) using a non-contact infrared thermometer (NCIT). We also measured the tympanic temperature using a tympanic thermometers (IRTT) and defined fever as a tympanic temperature ≥37.3°C.

**Results:** We enrolled a total of 528 participants including 261 indoor and 267 outdoor participants. We divided outdoor participants into four types according to their means of transportation to the hospital as walk, bicycle, electric vehicle, car, and inside the car. Under different circumstance, the mean difference ranged from −1.72 to −0.56°C in different groups for the forehead measurements, and −0.96 to −0.61°C for the wrist measurements. Both measurements had high fever screening abilities in inpatients (wrist: AUC 0.790; 95% CI: 0.725-0.854, *P* <0.001; forehead: AUC 0.816; 95% CI: 0.757-0.876, *P* <0.001). The cut-off value of wrist measurement for detecting tympanic temperature ≥37.3°C was 36.2°C with a 86.4% sensitivity and a 67.0% specificity, and the best threshold of forehead measurement was also 36.2°C with a 93.2% sensitivity and a 60.0% specificity.

**Conclusions:** Wrist measurement is more stable than forehead measurement under different circumstance. Both measurements have great fever screening abilities for indoor patients. The cut-off value of both measurements was 36.2°C. (ClinicalTrials.gov number: NCT04274621)

## Introduction

The outbreaks of 2019 novel coronavirus COVID-19 (previously known as 2019-nCoV) has attracted global attention, due to its strong transmission ability and certain fatality (1, 2). Some studies reported that fever, fatigue and dry cough are common symptoms of COVID-19 patients (3, 4), and 43.8% of the patients showed fever before admission with it largely being the first symptom (5). Therefore, temperature screening in the high-risk population is important for early identification of COVID-19 infection and thereby reducing the risk of cross infection.

During the epidemic, infrared tympanic thermometers (IRTT) and non-contact infrared thermometer (NCIT) are being applied to temperature screening in the general population (6). As a screening tool, it is quick for mass screening and allows a faster triage (7). However, we need to consume a lot of disposable plastic covers when we use IRTT. It may increase the financial burden in the widespread use of population screening. Furthermore, indirect contacts with infected individuals may increase the risk of cross infection. NCIT meets the clinical requirements for mass screening in terms of detection efficiency, safety and cost-performance. Besides, it takes less time than IRTT. Forehead is one of the key targets of thermography. However, forehead temperature is affected by physiological and environmental conditions (8). It should be measured in a relatively temperature-controlled environment. A previous study suggested to acclimate to the indoor temperature for at least 10 min for those who were exposed to the cold before taking body temperature readings (8). However, it is not practical for mass screening in winter during the outbreak of COVID-19.

Wrist temperature in this outbreak is under consideration. Before testing, they just need to roll up their sleeves at 10 cm above the palmar side of the wrist. Considering this area is covered with clothing, the wrist temperatures may keep stable. Previous study showed wearable devices (WD) on the wrist were applied in temperature monitoring in clinical practice (9). It brings a challenge whether it can be used as an accurate, safe and cost-effective screening tool in this outbreak.

In this study, we explored the accuracy and advantages of wrist temperature measurement in a real-life population in different environments and conditions. We aimed to find the thresholds of this key technique for diagnosis of fever. It may assist to improve the standardization of both practical use and performance, especially indispensable in the pandemic 2019-nCoV situation.

## Materials and Methods

### Study population

This was a prospective observational study in a real-life population. We consecutively enrolled a total of 572 participants at Ningbo First Hospital in China in this study (Figure 1). The exclusion criteria included: (i) Age ≤ 18 years (*n* = 6); (ii) Wearing hearing aid, or having a cerumen (*n* = 7); (iii) Participants with soft tissue infection or trauma (*n* =3); (iv) Missing data of wrist, forehead, and tympanic temperature (*n* = 4); and (v) Participants whose forehead temperature measurements showed “low” (*n* = 23). We finally enrolled 528 eligible participants for the final analysis, including 261 indoor and 267 outdoor participants. The 261 indoor participants were from the fever clinic and emergency department, and the 267 outdoor participants included patients and accompanying family members. The data of indoor participants were collected consecutively between February 14th and February 20th, 2020. The data of outdoor participants were collected on February 14th, 15th, 17th, 2020. Temperature readings were taken by trained and experienced nurses. Each participant was measured for wrist, forehead, and tympanic temperature twice. The temperatures were recorded by mean wrist temperature, forehead, and tympanic temperature, respectively. Data regarding age, gender, transportation, occupation, and temperature were recorded immediately by the nurse to pre-printed files.

**Figure 1.**
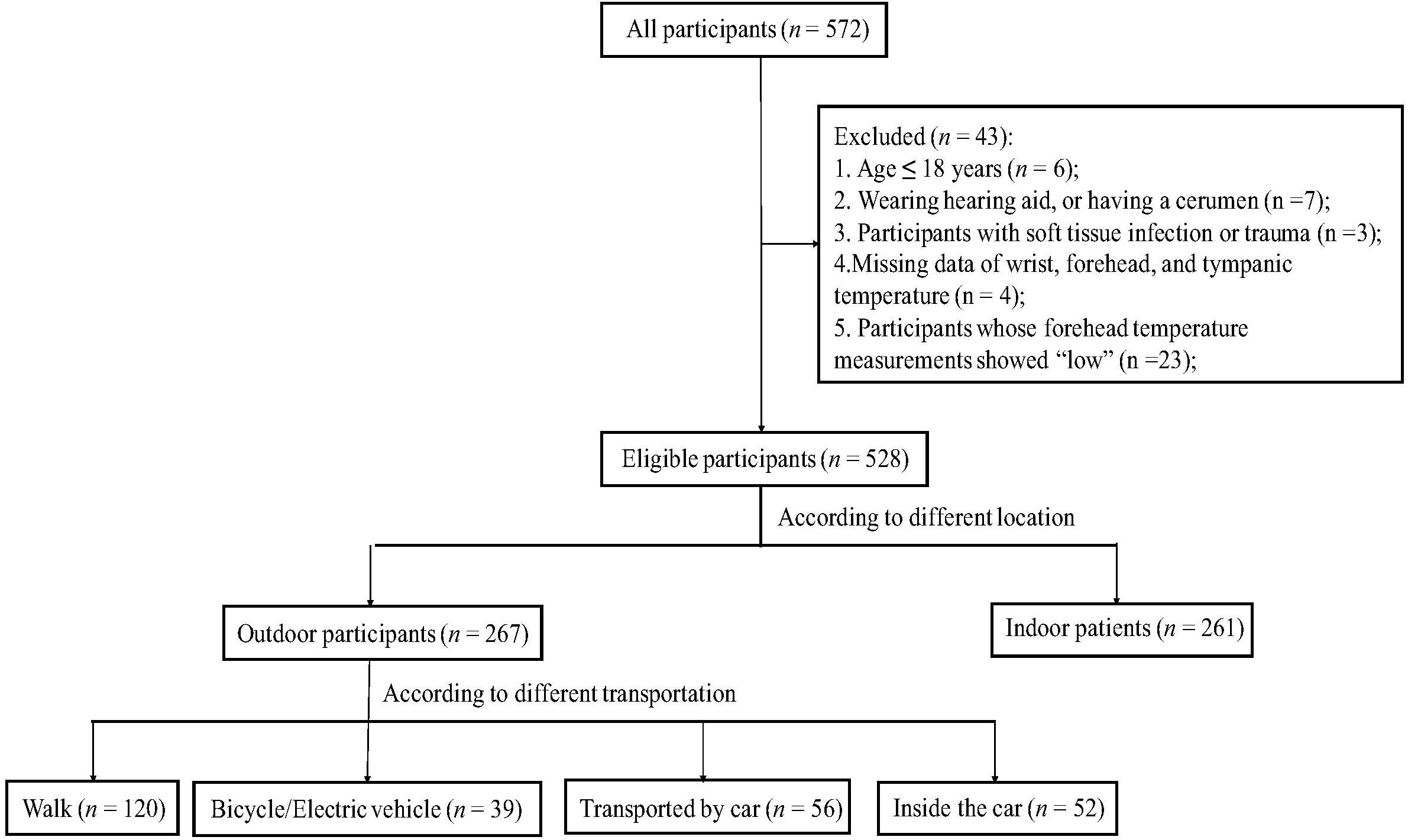
Flowchart of the study.

The study was approved by Ningbo First Hospital Ethics Committee. All participants were asked verbally. They gave their oral informed consent in this study. The study was registered in ClinicalTrials.gov with identifier number: NCT04274621.

### Assessment of environment

Indoor patients at the fever clinic and emergency department were those who has been indoors for at least a few minutes. The outdoor participants were divided into four type according to their means of transportation to the hospital as walk, bicycle/electric vehicle, car, and inside the car.

### Measurement of temperature

Tympanic temperature was measured using IRTT (Braun ThermoScan PRO 6000). Wrist and forehead temperature were measured using NCIT. The NCIT was ranged 32.0–42.9°C. The accuracy was ± 0.2°C. NCIT measurements were taken following the manufacturer’s instructions in the mid-forehead and a region at 10 cm above the palmar side of the wrist. After pulling the pinna backward, the nurse inserted IRTT into the external auditory meatus. The probe was held in the same position until the “beep” was heard. Temperature readings were taken by the same trained nurse in the following order: forehead, forehead (the second time), left wrist, right wrist, left tympanic, and right tympanic. The data were recorded by another researcher in pre-printed files. Tympanic membrane is in close proximity to the hypothalamus and the internal carotid artery (10). Thus, tympanic temperature is considered to directly reflect core temperature (11), and was defined as the gold standard in this study. These thermometers were stabilized before measurements. Calibration of thermometers were checked by the Quality and Technology Supervision Bureau, Ningbo, China. It was according to Calibration Specification of Infrared Thermometers for Measurement of Human Temperature (JJF1107-2003).

### Statistical analysis

Power calculation was performed for sample size. The following parameters were used: a power of 90%, an α-error level of 0.05, estimating a standard deviation of 1°C and a potential allowable error of 0.2°C. Considering a 10% possibility of dropouts and otherwise missing data, at least 293 subjects were needed in our study.

Continuous variables were expressed as mean ± standard deviation (SD), and categorical data in frequency and proportion. The agreements for each method versus tympanic temperature were analyzed by Bland–Altman analysis (12). It also showed three superimposed horizontal lines. Red dashed line highlighted mean bias among all the paired measurements. Black dashed line marked upper and lower 95% Limits of Agreement (LoA). A temperature deviation of 0.5°C was considered as clinically acceptable (13). A tympanic temperature of ≥ 37.3°C was defined as the cut-off point for fever. Statistical analyses were conducted using R version 3.5.1 (The R Foundation for Statistical Computing, Vienna, Austria).

## Results

### Participants

In this prospective observational study, a total of 528 participants were enrolled. Figure 1 summarizes characteristics of the participants. The mean age was 46.7 ± 16.4 years. 69.4% (*n* = 297) of participants were males, and 78.2% (*n* = 413) were patients (Table 1). Mean forehead, wrist, tympanic measurements were 35.6 ± 1.2°C, 35.7 ± 0.8°C, and 36.6 ± 0.6°C, respectively. There were 44 patients with fever in indoor patients. The data of outdoor participants were collected on February 14th, 15th, 17th, 2020. Mean weather temperatures were 13°C, 14°C, and 7°C, respectively.

**Table 1.** Demographic characteristics of the participants

| Variables | Total (*n* = 528) |
| --- | --- |
| Age, years | 46.7 ± 16.4 |
| Gender, male, *n* (%) | 297 (69.4%) |
| Environment |  |
| Indoor patients, *n* (%) | 261 (49.4%) |
| Walk, *n* (%) | 120 (22.7%) |
| Bicycle/Electric vehicle, *n* (%) | 39 (7.4%) |
| Transported by car, *n* (%) | 56 (10.6%) |
| Inside the car, *n* (%) | 52 (9.8%) |
| Patients or not |  |
| Yes, *n* (%) | 413 (78.2%) |
| Forehead temperature, °C | 35.6 ± 1.2 |
| Wrist temperature, °C | 35.7 ± 0.8 |
| Tympanic temperature, °C | 36.6 ± 0.6 |

### Bland-Altman comparison among the participants under different environment

Table 2 showed mean temperatures and Bland-Altman analysis among the participants under different environment. Compared with tympanic temperature as golden standard, the mean difference ranged from −1.72 to −0.56°C for the forehead measurement, and −0.96 to −0.61°C for the wrist measurement. We observed a lower variation in wrist than forehead temperature measurements.

**Table 2.** Bland-Altman comparison among the participants under different environment

| Environment | Method | Mean<br>temperature (°C) | Bland-Altman comparison (°C) |  |  |
| --- | --- | --- | --- | --- | --- |
|  |  |  | Mean difference | 95% prediction interval | Proportion of Differences within 0.5°C |
| Indoor patients | Tympanic | 36.8 | reference |  |  |
|  | Wrist | 35.8 | -0.96 | (-2.70—0.77) | 41.4% |
|  | Forehead | 36.2 | -0.56 | (-1.91—0.80) | 57.1% |
| Walk | Tympanic | 36.3 | reference |  |  |
|  | Wrist | 35.4 | -0.86 | (-2.05—0.34) | 72.5% |
|  | Forehead | 34.6 | -1.72 | (-4.07—0.64) | 22.5% |
| Bicycle/Electric<br>vehicle | Tympanic | 36.0 | reference |  |  |
|  | Wrist | 35.5 | -0.61 | (-2.14—0.93) | 56.4% |
|  | Forehead | 34.6 | -1.49 | (-3.82—0.84) | 48.7% |
| Transported by<br>car | Tympanic | 36.6 | reference |  |  |
|  | Wrist | 35.7 | -0.93 | (-1.43—-0.44) | 91.1% |
|  | Forehead | 35.4 | -0.92 | (-1.47—-0.36) | 85.7% |
| Inside the car | Tympanic | 36.7 | reference |  |  |
|  | Wrist | 35.8 | -0.85 | (-1.54—-0.15) | 94.2% |
|  | Forehead | 35.8 | -1.13 | (-2.41—0.16) | 80.8% |

Outdoor participants were divided into four types as walk, bicycle or electric vehicle, car, and inside the car. For those who walked, the agreement limits for wrist and tympanic was between −2.05 and 0.34°C; −4.07 and 0.64°C for forehead and tympanic (Figure 2A, B). For those who used bicycle or electric vehicle, the agreement limits for wrist and tympanic was between −2.14 and 0.93°C; −3.82 and 0.84°C for forehead and tympanic (Figure 2C, D). For those who were transported by car, the agreement limits for wrist and tympanic was between −1.43 and −0.44°C; −1.47 and −0.36°C for forehead and tympanic (Figure 2E, F). For those who were inside the car, the agreement limits for wrist and tympanic was between −1.54 and −0.15°C; −2.41 and 0.16°C for forehead and tympanic (Figure 2G, H). It highlighted that wrist temperature had narrower 95% limits of agreement than forehead. Wrist measurements had the higher percentage of differences falling within ± 0.5°C than forehead measurements in these four types.

**Figure 2.**
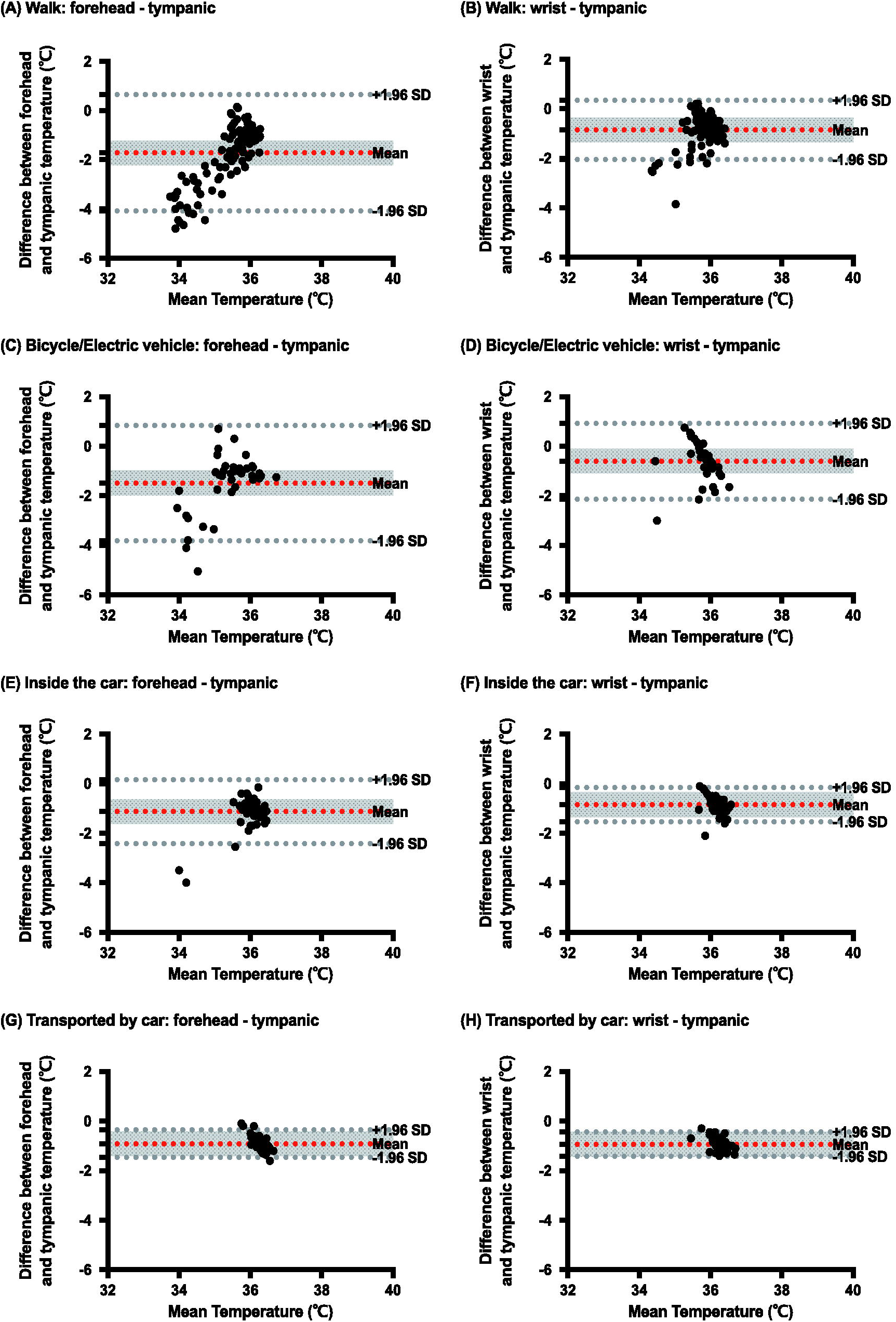
Bland-Altman comparison between each method and tympanic temperature. X axis is the mean temperature of each method and tympanic. Y axis is the difference of each method and tympanic. Red dashed line showed mean bias. Black dashed lines showed 95% limits of agreement. (A), (B) for those who walked; (C), (D) for those who used bicycle/electric vehicle; (E), (F) for those who were transported by car; (G), (H) for those who were inside the car

For indoor patients, the agreement limits for wrist and tympanic was between −2.70 and −0.77°C; −1.91 and 0.80°C for forehead and tympanic (Figure 3). 57.1% of forehead values were included within ± 0.5°C, followed by wrist values (41.4%). We also explore the agreement of left and right wrists (Figure S1). The mean bias is 0.00. The agreement limits for wrist and tympanic was between −0.74 and 0.74°C. It showed good agreement between right and left wrists.

**Figure 3.**
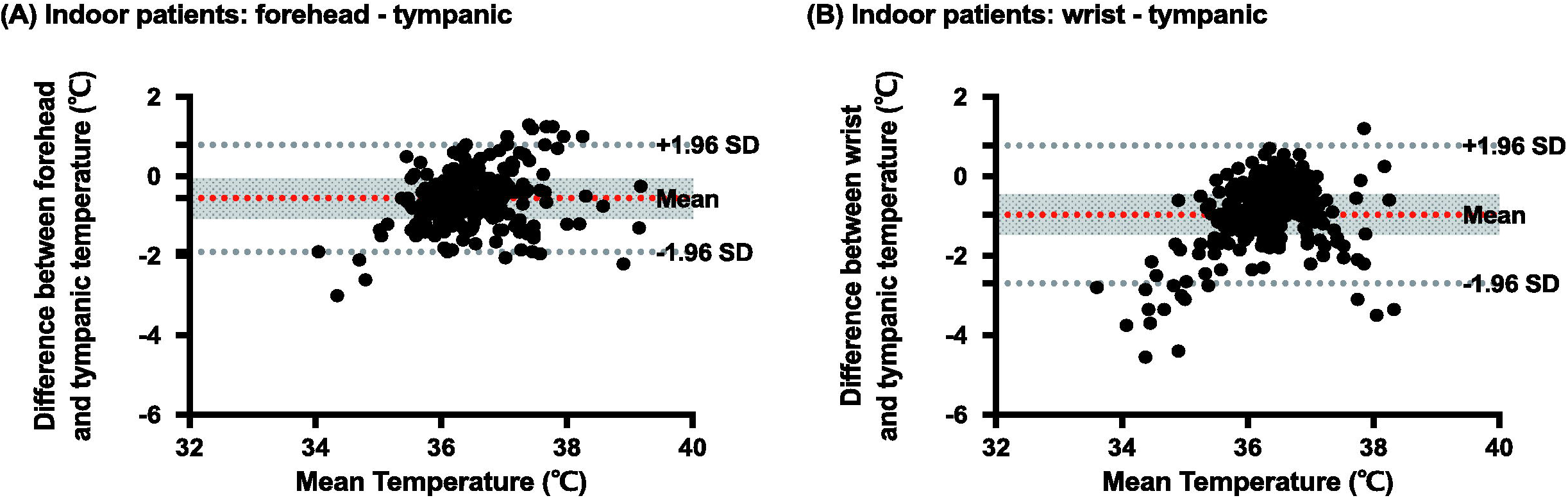
Bland-Altman comparison between each method and tympanic temperature for indoor patients. X axis is the mean temperature of each method and tympanic. Y axis is the difference of each method and tympanic. Red dashed line showed mean bias. Black dashed lines showed 95% limits of agreement.

### The receiver–operator characteristic (ROC) curves for detection of fever

We performed a ROC curves in indoor patients for detecting tympanic temperature ≥37.3°C. Figure 4 shows the comparison between wrist and forehead measurements for detection of fever. Both measurement had significantly great abilities of screening patients for fever (wrist: AUC 0.790; 95% CI: 0.725–0.854, *P* <0.001; forehead: AUC 0.816; 95% CI: 0.757–0.876, *P* <0.0001). The cut-off value of wrist measurement for detecting tympanic temperature ≥37.3°C was 36.2°3 with a 86.4% sensitivity and a 67.0% specificity. And the best threshold of forehead measurement was also 36.2°6 with a 93.2% sensitivity and a 60.0% specificity.

**Figure 4.**
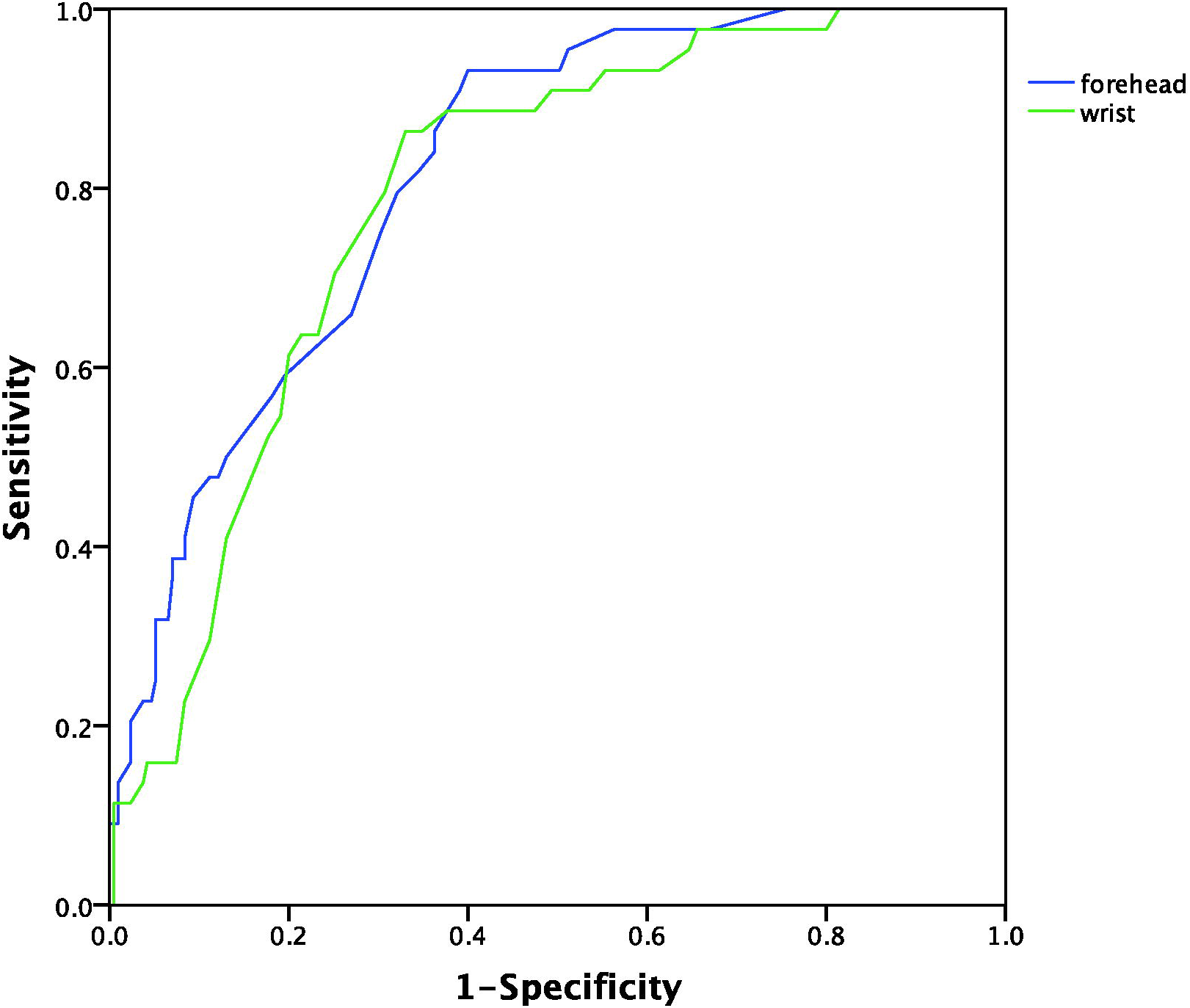
The receiver–operator characteristic (ROC) curves for detection of fever.

## Discussion

In this prospective real-world study, we found that wrist temperature measurement is more stable than forehead using NCIT under different circumstances in outdoor participants. Both measurement had significantly great abilities of screening patients for fever in indoor patients. The cut-off value for wrist and forehead temperature were both 36.2°C. They showed good sensitivity. It may assist for fever screening in the population, especially in the outbreak of 2019 Novel Coronavirus (COVID-19). To our knowledge, this study was the first to explore the reliability and validity of wrist and forehead temperature measurement in mass screening.

Previous studies showed that axilla, rectal temperature were the gold standards in clinical practice (14, 15). However, it was impractical for the large-scale screening. Timesaving and less invasive tools were needed. IRTT and NCIT are being applied in the general population during the epidemic. A lot of disposable plastic covers were consumed, which may increase the financial burden. In China, it cost 1–2 RMB (about 0.2 dollars) for per disposable plastic cover. Besides, indirect contacts increased the risk of cross infection. Forehead temperature was used for the widespread use of population screening using NCIT. However, it can be affected by a person’s physiological and environmental conditions (8, 16). The forehead temperature value of 23 participants showed “low” in our study. This all happened on the same day (February 17th, 2020) with an outside temperature of 7°C. Thus, we chose wrist temperature as an alternative, especially in the winter when mass screening is needed. Wrist measurement indicated peripheral temperature at 10 cm above the palmar side of the wrist. It was within our expectation that wrist measurement readings attained was lower than tympanic route. However, this area was covered by clothing all the time. It was less influenced by environmental conditions. Our study showed it was more stable for participants under different circumstance than forehead measurement. It is important for mass screen in the open air during the Outbreak of COVID-19. The ROC curves showed wrist and forehead measurement had significantly great abilities of screening patients for fever. The cut-off value of both measurement was 36.2°C. It can be applied in clinical practice and assist to improve the standardization of both practical use and performance.

The strengths of this study included its large sample size, and prospective design in the real-world setting. There were several limitations. First, it is difficult to quantify the physiological and environmental conditions. Second, only one brand of thermometer was enrolled in this study. It was uncertain that it could be generalized to all brands of thermometers in the market.

In conclusion, this study confirmed wrist measurement was more stable for participants under different circumstance than forehead measurement. Both measurement had significantly great fever screening abilities for indoor patients, and the cut-off value of both measurements for fever was 36.2°C. Further studies are needed to explore the validity and accuracy of wrist temperature.

## Data Availability

The data used to support the findings of this study are available from the corresponding author (Xiaoming Chen).

## Abbreviations

2019-nCoV: 2019 Novel Coronavirus
NCIT: Non-contact infrared thermometer
IRTT: infrared tympanic thermometers
ROC: receiver–operator characteristic

## Guarantor of the article

Xiaoming Chen

## Specific author contributions

Study concept and design: Ge Chen, Lei Xu, Xueqin Chen, and Xiaoming Chen; Acquisition of data: Peijun Zheng, Xiaqing Hu, Guangli Dai, Lei Xu and Hongpeng Lu; Analysis and interpretation of data: Jiarong Xie and Ge Chen. Drafting of the manuscript: Jiarong Xie and Ge Chen. Study supervision: Lei Xu, Xueqin Chen, and Xiaoming Chen.

## Author Approval

all authors have seen and approved the manuscript.

## Acknowledgment

The authors thank Jihong Zhang for sorting of data; Ximing Jiang, Ying Xin for acquisition and sorting of data.

